# Is the spread of COVID-19 across countries influenced by environmental, economic and social factors?

**DOI:** 10.1101/2020.04.08.20058164

**Authors:** Mohammad Alamgir Hossain

## Abstract

The SARS-CoV-2 virus, emerged from Wuhan, China is spreading all over the world in an unprecedented manner, causing millions of infections and thousands of deaths. However, the spread of the disease across countries and regions are not even. Why some countries and regions are more affected than some other countries and regions? We employ simple statistical methods to investigate any linkage between the severity of the disease and the environmental, economic and social factors of countries. The estimation results indicate that the number of confirmed cases of Coronavirus infection is higher in countries with lower yearly average temperatures, higher economic openness and stronger political democracy. However, findings of this analysis should be interpreted carefully keeping in mind the fact that statistical relations do not necessarily imply causation. Only clinical experiments with medical expertise can confirm how the virus behaves in the environment.

## Introduction

The ongoing global Coronavirus pandemic started all on a sudden and has posed serious threat for the existence of normal human life. The novel virus, named afterward as SARS-CoV-2, belongs to highly pathogenic Coronavirus family and causes severe acute respiratory disease, COVID-19 leading even to death for many cases ^1^. The speed of human-to-human transmission of the pathogen is unprecedented. It has spread to most of the countries of the world within 4 months of its first known transmission to human and the number of infected people as well as the number of deaths caused by the virus is increasing every day till date. The first human case of present outbreak of Coronavirus is identified in Wuhan, the capital of Hubei province of China, in December, 2019^1,2^. Though the virus is spreading many countries, the speed and severity of spread vary country to country. Europe is badly affected than Asia, Africa and South America. Though South Korea, Thailand, Japan and Singapore were affected immediately after China, wide-spread infections were observed in Italy and Spain before those countries. Despite having political and economic connection and national borders with China, Myanmar and North Korea report very few cases of infection. Other European countries and USA were also hit within a very short span of time. Nevertheless, other Asian, African and South American countries are less affected till the beginning of April 2020. The uneven transmission mechanism to countries raises the question- what factors do influence the pathogen to spread? Is the virus attacking countries with lower temperature, or higher precipitation, or higher openness? Biological explanation can answer many questions related to transmission mechanism of a pathogen. Apart from biological properties, it is susceptible that few environmental, social and economic elements can act as possible catalysts for the spread of novel Coronavirus. First, connection with China is likely to be an important factor for rapid transmission of the disease at the initial stage of the pandemic. As the virus continues to spread many countries, any connection with badly effected countries may equally responsible for the transmission. In other words, level of international connection of a country is probably playing a vital role in country-to-country transmission of the virus. International trade, alternatively economic openness can be treated as a good measure of international connection of a country. Thus, economic openness is likely to play an important role in the rate of spread of the disease in a country. If a country is more open, the virus will find it easier to transmit from human to human by crossing national borders. Once the virus is in, the rate of spread within a country will depend on several factors, some of which are difficult to identify, some though can be pointed out by educated guess. Population density, level of urbanization, social cohesiveness and weather conditions can be primarily identified as influential factors of human-to-human transmission of a pathogen. Viability of infectious viruses is found being affected markedly by environmental factors like temperature and humidity ^3^. Social, economic and environmental changes around the world are also believed to increase the occurrences of infectious diseases in the world, especially in developing countries ^4^. In case of SARS-CoV-2 pandemic, many countries responded immediately and are imposing partial or full lockdown. Measures taken by governments and response rate of citizens at the initial stage of the outbreak at any locality are likely to impact significantly on the spread ^5^. Furthermore, published number of confirmed cases are not completely reliable for some reasons. There is some evidence that fear of economic penalties sometimes lead authorities to under-report epidemics ^4^. Countries of autocratic rulers or non-democratic government are reluctant to reveal exact or approximate number of cases as it may raise questions about their governance. Moreover, social cohesiveness is likely to be higher in free societies leading to possibilities of more infections of a highly pathogenic disease. Though, democratization does not necessarily mean social cohesiveness or integrity, it can be used as a proxy to assess how much the citizens of a society are involved in social activities. If social activities are higher, the chance of more infections are higher. Thus, the level of democratization is a possible inducer of the number of cases of infection.

Underlying factors influencing the present spread mechanism of SARS-CoV-2 across countries are not well-known yet. Researchers around the world are contributing to better understand the virus that may save lives.

Present study is motivated to find any possible linkage between world-wide uneven spread of COVID-19 and environmental, social and economic characteristics of countries by applying statistical methods. Though statistical relationship does not mean causation, they are at least indicative to a true relationship. We use least squares method to investigate whether the transmission of COVID-19 across countries is influenced by few environmental, social and economic variables.

## Trends of COVID-19 Spread

On 12 January 2020 the World Health Organization (WHO) announced that a novel Coronavirus is infecting clusters of people in Wuhan, China from December, 2019 causing respiratory syndromes ^1^. On 13 January, a case was reported in Thailand, the first outside of China, in a woman who had arrived from Wuhan^2^. The second case outside of China was found on 15 January in a man in Japan who visited Wuhan. The case of infection in USA was declared on 21 January. On 24^th^ January, the WHO notified first confirmed case of human-to-human transmission of the virus in Vietnam ^2^. The WHO declared a global health emergency on January 30. Two months afterward was the time of Corona. The number of infected people was rising rapidly. Due to time variation across countries, figures on infection and death by Coronavirus on a specific day differs slightly by sources. As published on the website of ECDC, on 05 April 2020 world-wide total confirmed cases of infection reaches at 1174652 and world-wide total number of deaths reaches at 64400. The global number of infections of Coronavirus doubles approximately in 8 days and the number of deaths in 7 days. After China, the worst affected regions are Europe and USA. Europe is expecting a downturn after a long upward rise, while USA is supposed to experience the rise further. Many third world countries of Asia and Africa have just started experiencing of COVID-19, with huge anxiety about possible fatality that may happen. The trends of China, Europe and USA in the figure-1 indicate a similar steepness, whereas they differ in time of spread. The graph of India, another populous country, now at its early stage of infection, exhibits almost similar steepness.

**Figure-1:**
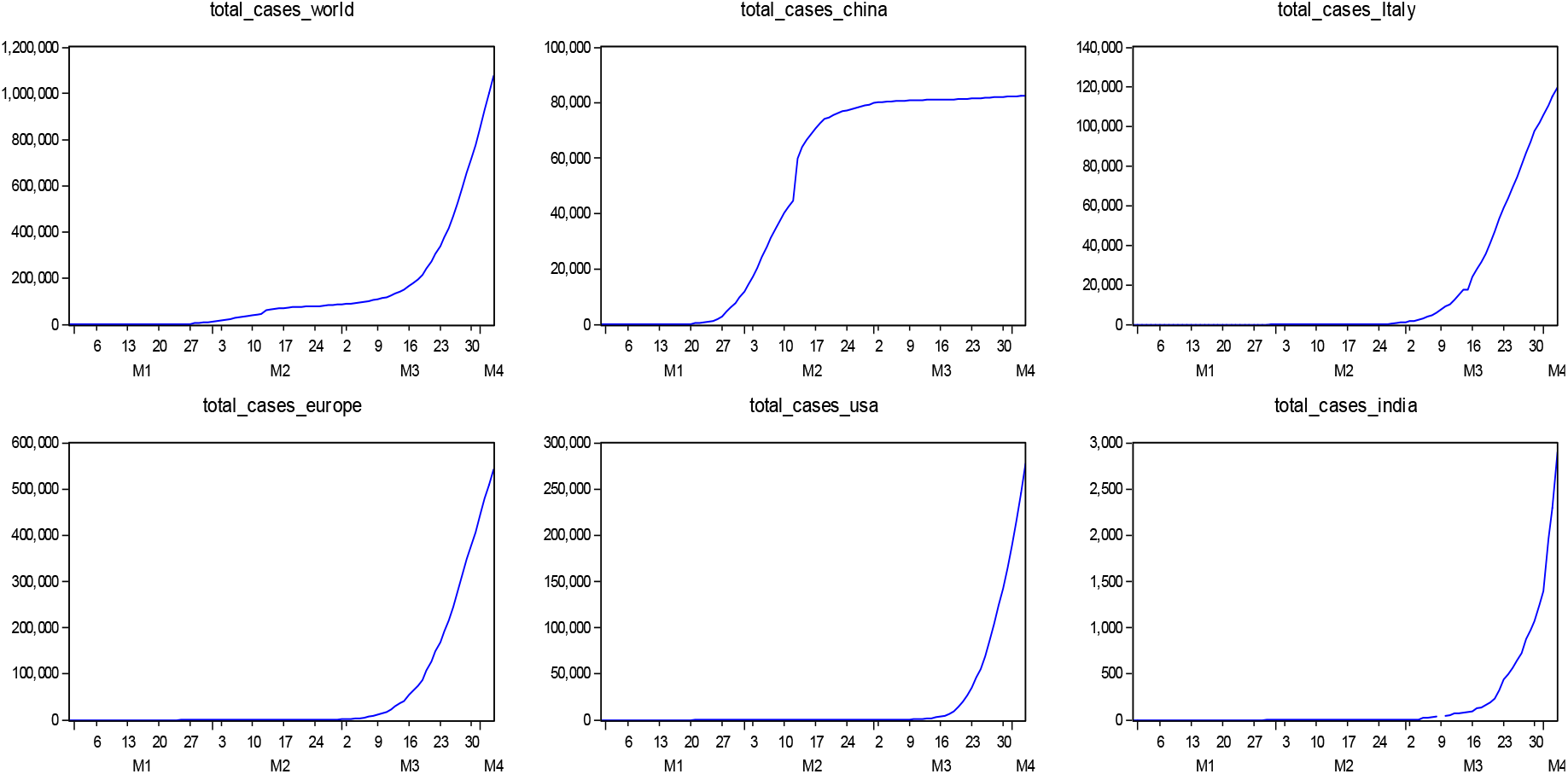
Trends in total cases of infection.

**Figure-2:**
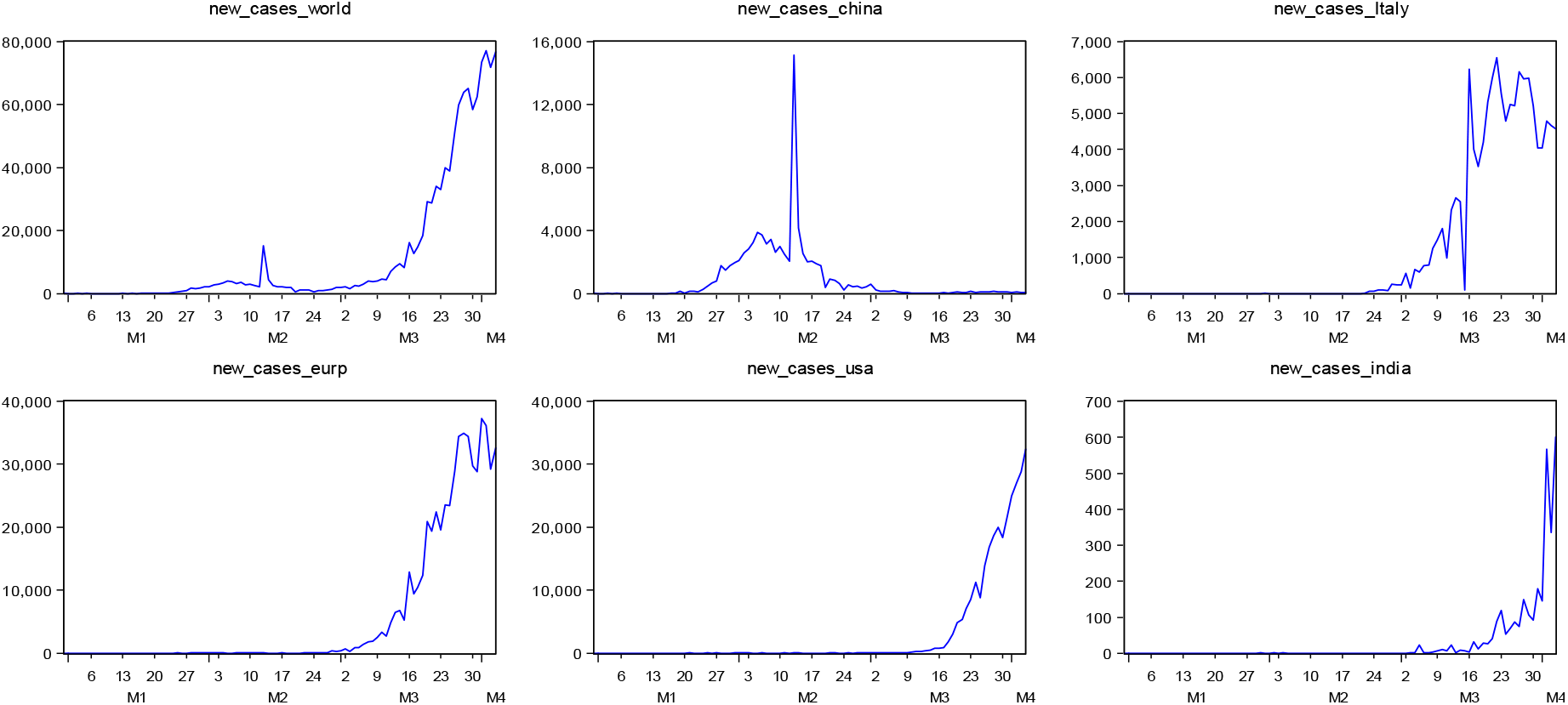
Trends in daily new cases.

Patterns of daily new cases for Italy, Europe, USA and India are almost similar showing a very steep upward trend, though fluctuations are more obvious in case of Italy. Fluctuations in the trends of daily new cases are probably highly influenced by governments’ interventions. China reached its peak on 13 February 2020 with 15141 new cases on that day. On 12 February number of new cases was 2028, and on 14 February the number of new cases was 4156 in China.

This sharp rise and down of 13 February in China are surprising and deserves better explanation. Few big spikes are also visible in cases of Italy and India.

On 16 March Italy got new cases of 6230, whereas the number was only 90 on the previous day. On that day, the trend line of Italy got structural upward shift. In case of USA, the upward rise of daily new cases is comparatively smoother. India, a late affected country, is exhibiting upward rise in daily new cases with more fluctuations. In very recent days, India experiencing infections of Coronavirus with an increasing growth rate.

## Determinants of Infectious Disease Spread

Studies on transmission patterns of infectious diseases including viruses causing respiratory syndromes, like SARS Coronavirus are available in the literature, though further researches are required. Rottier & Ince (2003) describes that transmission cycle of a pathogen depends, in general, on the pathogen itself, host and/or intermediate host, the environment and the availability of susceptible person or animal. Precipitation, humidity, temperature, and airflow can be determinants of virus infection and transmission ^7^.

The biological and transmission mechanisms of SARS-CoV-2 are not well-known yet ^8^ and under extensive investigation by researchers throughout the world. A very recent study claims that high temperature and high relative humidity reduce reproductive number, R_0_ of Coronavirus significantly and consequently lessen the transmission of the disease ^9^. Human-to-human transmission of SARS Coronavirus is likely to be influenced also by airflow and ventilation as indicative in a research conducted in Hong Kong ^7^.

According to Guo et al (2020) human-to-human transmission of SARS-CoV-2 occurs mainly through a close contact with patients of COVID-19 or incubation carriers. In a groundbreaking work, published on 30 January 2020, Riou and Althaus (2020) apprehended that the spread of the novel virus will depend on two key properties. First, the reproduction number, R_0_ (average number of secondary cases by an infectious index case) value of which is estimated as 2.2, greater than the threshold level, 1. Second, the dispersion parameter of secondary cases, k whose value is liable for super-spreading. A low value of k would lead a steady growth of the epidemic. Though the outbreak appears to have started from a single or multiple zoonotic transmission in Wuhan ^10^, the spread outside of China is clearly Human-to-human transmission. Human-to-human transmission may be direct or indirect. Direct Human-to-human transmission occurs because of close contact with a patient. However, the virus can stay in the environment for sometimes and can infect a susceptible person if get contacted. This indirect Human-to-human transmission is likely to depend on many factors including pathogenic characteristics and environmental factors.

This study aims to explore the effects of few environmental, social and economic characteristics of countries on country-wise number of infection cases per one million people. We hypothesize that number of infection cases per one million people of a country depend on yearly average temperature, yearly average precipitation, economic openness, level of democracy, and on population density of the country.

How long the virus can stay alive in the environment without any receptor will depend on temperature and precipitation influencing the rate of indirect human-to-human transmission. On the other hand, number of cases of infection in a country will usually be accelerated by the number of imported cases from abroad, either from China or via other country. If more people enter a country as a vector of the pathogen, the secondary number of cases must be higher inflating the total number of cases. The number of imported infection cases is likely to increase if the country is involved in higher degree of international trade indicating economic openness as a determining factor of the ongoing pandemic. Many social conditions of a country may influence the shape of the spread. A cohesive society where people are involved in more festivals or religion gatherings is more likely to have more cases. Many other cultural issues may also have connection with the disease. We consider the level of democratization as a proxy variable for social cohesiveness. Population density is also likely to have positive association with the spread of COVID-19.

## Data sources and characteristics

Data on Coronavirus infection cases along with population in 2018 are collected from the website of European Centre for Disease Prevention and Control (ECDC). Average yearly temperature and average precipitation data were collected from meteoblue.com and indexmundi.com respectively. Openness data as defined by ratio of international trade to GDP were downloaded from Databank of the World Bank. The scores of democracy index 2019, compiled by Economist Intelligence Unit, UK, is collected from Wikipedia. As the sources are various in type, we cannot include all countries infected by COVID-19 in our analysis due to unavailability of all data for few countries. Total 163 countries are included in this analysis.

Total number of cases of Coronavirus infection by countries is time variant. The number accumulates over time. Nonetheless, total cases of infection in a specific country on a specific day (Y) is a fixed number. Thus, the variable, total cases of infection by countries has both time series and cross-section elements. We collect data on total infection cases by countries from 31 December 2019 to 03 April 2020. Total cases of infection are converted to cases per one million population to capture the population effect. Cases of infection per one million people on 03 April 2020 by countries is denoted by Y and used as the dependent variable in our experimentation. Values of this variable obviously depend on its previous values. Consequently, Y is simultaneously a cross-section and time series variable. We consider country level yearly average temperature and precipitation as environmental variables. On the other hand, the economic and social variables, viz. openness, democracy index and population density are also measured on yearly basis. Accordingly, the explanatory variables we are dealing with are static in nature.

## Modeling Approach

Transmission mechanism of a pathogen is mainly characterized by biological properties. Total number of cases of an infectious disease within a country and at a point of time is determined by a lot of biological and non-biological factors. Thus, we can presume that total number of confirmed cases across countries is generated by a stochastic process. However, the number that observed at a point of time is basically a realization of the process through which data is generated ^11^. The realized value is a random selection from a range of possible values that could be generated by the interaction of all factors. Though it is not easy to reveal the true characteristics of the underlying data generation process, the realized or observed values can be utilized to draw inferences about the process. As a stochastic process the important determinant of the number of cases of a day is its own past values, implying that Y_t_, the number of cases on a day, depend on the past values, i.e. Y_t-1_, Y_t-2_, Y_t-3_ etc. Hence, an autoregressive model may be suitable to approximate the data generation process of the cases of infection on a day. Nevertheless, our aim in this study is not to find the data generation process of total number of cases of a country on a specific day. Rather, our objective is to examine data to investigate any possible linkage between country level severities of infection with few selected environmental, economic and social variables. For this purpose, we first simply regress total number of cases of infection per one million people by countries reported on a recent day (03 April 2020) on our selected explanatory variables. Specifically, we estimate the following regression-

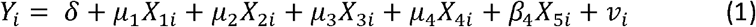

Y is the total number of cases of confirmed infection per one million people in a country on a day (03 April 2020), X_1_ is yearly average temperature of countries, X_2_ is yearly average precipitation of countries, X_3_ is openness measured by international trade as a percentage of GDP of countries, X_4_ is democracy index of countries and X_5_ is population density of countries in 2018.

We also presume that total number of cases of infection in a country on a specific day depends, apart from environmental, social and economic conditions, on the previous number of infection cases. Thus, in addition of model (1) we aim to fit a regression model of the following type-

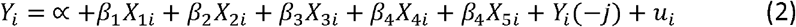

where, j means period of lag. The value of j should be selected based on best fit of the model as the timing and duration of viral shedding after SARS-CoV-2 infection is unknown ^12^. If model (2) represents a true relationship, then it will suffer from multicollinearity as lagged Y must have high correlation with time invariant explanatory variables included in the regression equation. In the presence of multicollinearity, the estimated coefficients will be biased. To get rid of the multicollinearity problem, we first regress the lagged Y on rest of the explanatory variables and get the residual series. Henceforth, we run the following regression-

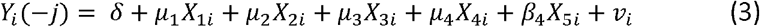

From the estimated values of model (3), we get-

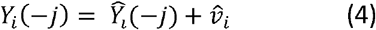

where 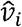 is the residual series. Following the properties of Classical Linear Regression Model, we can deduce that 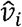 of the estimated model (3) is un-correlated with X’s and can be used as a proxy variable of *Y*_*i*_ (-*j*) in model (2). Consequently, model (2) has converted to the following estimable one-

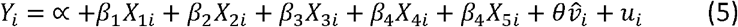

This model can be used to draw inferences about any linear relationship between Y and the X’s holding the effect of past values on Y. We use Excel and STATA package for manipulation and transformation of data. For estimation purpose we use EVIEWS.

## Estimation Results and Discussion

We apply Least Squares method on model (1) and find that precipitation and population density have no significant effect on the number of infection cases per one million people (Y). Those variables are then excluded, and the model is re-estimated. In the re-estimated model, the variables average temperature, openness and democracy appear as highly significant. We made some experimentations with lags 1 to 9 to estimate model (5). In each case we first estimate model (3) and obtain the residuals, then estimate model (5) using the estimated residuals obtained from estimated model (3). As the number of infections are still increasing day by day in almost all countries, the estimated model fits better with lower lag. We drop density and precipitation from our experimentations as they frequently appear insignificant. In all estimated model yearly average temperature, openness and democracy appear as highly significant. Estimation results of three regression models with no lag, lag 1 and lag 9 are presented in Table-1.

**Table 1:** Estimation Results.

| Dependent Var.:<br>Cases of infection per Mil. On<br>03 April | Model 1<br>(no lag) | Model 2<br>(period of lag=1) | Model 3<br>(period of lag=9) |
| --- | --- | --- | --- |
| Temperature | -16.55022<br>(0.0064) | -16.55022<br>(0.0000) | -16.55022<br>(0.0000) |
| Openness | 2.687991<br>(0.0004) | 2.687991<br>(0.0000) | 2.687991<br>(0.0000) |
| Democracy | 86.76467<br>(0.0001) | 86.76467<br>(0.0000) | 86.76467<br>(0.0000) |
| $\hat{v}_i$ | - | 1.146961<br>(0.0000) | 2.347331<br>(0.0000) |
| C | -181.5641<br>(0.0000) | -181.5641<br>(0.0000) | -181.5641<br>(0.0000) |
| **Fit Measures** |  |  |  |
| $\bar{R}^2$ | 0.312467 | 0.998798 | 0.953022 |
| SER | 491.6117 | 20.55306 | 128.5060 |
| Akaiki info | 15.26161 | 8.919205 | 12.58514 |
| **Diagnostics** |  |  |  |
| BG LM | 2.658312<br>(0.26) | 0.395208<br>(0.82<sup>*</sup>) | 2.535723<br>(0.28) |
| LMH | 0.2647<br>(0.00) | 70.64418<br>(0.00) | 104.4686<br>(0.00) |
| Ramsey's RESET LR | 54.23739<br>(0.00) | 2.570001<br>(0.28<sup>*</sup>) | 0.714289<br>(0.70<sup>*</sup>) |
| JB | 1490.72 | 427.79<br>(0.00) | 708.13<br>(0.00) |

Note: Figures in the parenthesis below the coefficients and the statistics are the p-values. $\bar{R}^2$ is adjusted coefficient of determination. SER is the regression standard error. BG LM means Breusch-Godfrey Lagrange Multiplier test for serial correlation, LMH is the White's LM test for heteroscedasticity, JB is Jarque-Bera statistic.

All the estimated coefficients in three reported models are significant with expected signs. Values of estimated coefficients of yearly average temperature, economic openness and democracy index remain same in all models pointing at high stability of estimation results. The sign of temperature appears negative implying that warmer countries are less infected by the Coronavirus. This is an indication of a relationship between environmental factors and the transmission mechanism of SARS-CoV-2. Nevertheless, only clinical experiments can confirm the relationship. We did not find any relationship of Coronavirus infection with yearly average precipitation.

Positive sign of openness suggests that the more a country is involved in international trade, i.e. export and import, the more it is vulnerable to Coronavirus infection. This finding is plausible as more international trade bring forth more interaction with overseas people and pave the way for the viral infection.

In the same manner, the positive sign of democracy index indicates that more democratic countries are affected more by the disease. This is may be the result of the fact that the democracy index may imply social cohesiveness or social integration. People are likely to be more involved in social activities and festivals in a country which is more democratic than a country ruled by an autocrat. Moreover, autocratic rulers are generally reluctant to publish the actual figure of fatalities and try to undermine it in any crisis.

The estimated models are not free from any misspecifications. As the diagnostic tests indicate, the estimated models are suffering from heteroscedasticity and violate the normality assumption. However, the models are free from autocorrelation (except model 1) as indicated by Breusch-Godfrey LM test statistic and high multicollinearity as indicated by high 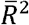 and significant t-ratios for all explanatory variables. Consequently, the estimated coefficients are unbiased, though do not possess minimum variances. Despite some deficiency, the estimated models admit that COVID-19 is likely to spread more countries with low yearly average temperature, higher international trade and higher level of democracy.

## Conclusion

In this study we employ Least Squares method and provide statistical evidences that yearly average temperature, economic openness and political democracy level of countries have significant effect on the spread of SARS-CoV-2 across countries. Data on 163 infected countries is analyzed latest on 03 April 2020 which reveals that countries with higher average temperature, lower international trade and weaker democracy tend to experience comparatively smaller number of cases of infection per one million people. Inclusion of more social and economic variables, for instance, level of urbanization, level of globalization, trade share with China etc. in the regression model may increase model validity. As the number of cases of infection accumulates over time for each country, panel data analysis including more time variant relevant variables may produce interesting outcomes. Government interventions are likely to influence the spread mechanism of Coronavirus and thus success of those interventions should be explored by further research.

The results of this analysis should be interpreted cautiously keeping in mind the fact that statistical relations do not necessarily imply causation. Only clinical experiments with medical expertise can confirm how the virus behaves in the environment.

## Data Availability

Secondary data is used in the research and sources are mentioned in the text.

## References

1. Guo Y-R, Cao Q-D, Hong Z-S, et al. The origin, transmission and clinical therapies on coronavirus disease 2019 (COVID-19) outbreak –an update on the status. Mil Med Res. 2020;7(1):1–10. doi:10.1186/s40779-020-00240-0

2. World Health Organization (WHO). Novel Coronavirus (2019-nCoV) Situation Report - 1 21 January 2020. WHO Bull. 2020;(JANUARY):1–7.

3. Park JE, Son WS, Ryu Y, Choi SB, Kwon O, Ahn I. Effects of temperature, humidity, and diurnal temperature range on influenza incidence in a temperate region. Influenza Other Respi Viruses. 2020;14(1):11–18. doi:10.1111/irv.12682

4. Frenk J, Gómez-Dantés O, Knaul FM. Globalization and infectious diseases. Infect Dis Clin North Am. 2011;25(3):593–599. doi:10.1016/j.idc.2011.05.003

5. Fang Y, Zhang S, Yu Z, Wang H, Deng Q. Shenzhen ‘Experience on Containing 2019 Novel Infected Pneumonia Transmission. 2020:2–4.

6. Rottier E, Ince M. Disease and disease transmission. Control Prev Dis role water Environ Sanit Interv. 2003:7–27. http://ec.europa.eu/echo/files/evaluation/watsan2005/annex_files/WEDC/diseases/diseases.htm.

7. Pica N, Bouvier NM. Environmental factors affecting the transmission of respiratory viruses. Curr Opin Virol. 2012;2(1):90–95. doi:10.1016/j.coviro.2011.12.003

8. Lau H, Khosrawipour V, Kocbach P, et al. Internationally lost COVID-19 cases. J Microbiol Immunol Infect. 2020;(xxxx):1–5. doi:10.1016/j.jmii.2020.03.013

9. Wang J, Tang K, Feng K, Lv W. High Temperature and High Humidity Reduce the Transmission of COVID-19. SSRN Electron J. 2020;(61572059):1–19. doi:10.2139/ssrn.3551767

10. Riou J, Althaus CL. Pattern of early human-to-human transmission of Wuhan 2019 novel coronavirus (2019-nCoV), December 2019 to January 2020. Euro Surveill. 2020;25(4):1–5. doi:10.2807/1560-7917.ES.2020.25.4.2000058

11. Gujarati DN, Porter DC. Basic Econometrics. 5th ed.New York: McGraw-Hill; 2009.

12. Ghinai I, McPherson TD, Hunter JC, et al. First known person-to-person transmission of severe acute respiratory syndrome coronavirus 2 (SARS-CoV-2) in the USA. Lancet. 2020;395(10230):1137–1144. doi:10.1016/S0140-6736(20)30607-3

